# Impact of SARS-CoV-2 variants on inpatient clinical outcome

**DOI:** 10.1101/2022.02.02.22270337

**Authors:** Matthew L. Robinson, C. Paul Morris, Josh Betz, Yifan Zhang, Robert Bollinger, Natalie Wang, David R Thiemann, Amary Fall, Raghda E. Eldesouki, Julie M. Norton, David C. Gaston, Michael Forman, Chun Huai Luo, Scott L. Zeger, Amita Gupta, Brian T. Garibaldi, Heba H. Mostafa

## Abstract

**Background:** Prior observation has shown differences in COVID-19 hospitalization rates between SARS-CoV-2 variants, but limited information describes differences in hospitalization outcomes.

**Methods:** Patients admitted to 5 hospitals with COVID-19 were included if they had hypoxia, tachypnea, tachycardia, or fever, and data to describe SARS-CoV-2 variant, either from whole genome sequencing, or inference when local surveillance showed ≥95% dominance of a single variant. The average effect of SARS-CoV-2 variant on 14-day risk of severe disease, defined by need for advanced respiratory support, or death was evaluated using models weighted on propensity scores derived from baseline clinical features.

**Results:** Severe disease or death within 14 days occurred for 950 of 3,365 (28%) unvaccinated patients and 178 of 808 (22%) patients with history of vaccination or prior COVID-19. Among unvaccinated patients, the relative risk of 14-day severe disease or death for Delta variant compared to ancestral lineages was 1.34 (95% confidence interval [CI] 1.13-1.55). Compared to Delta variant, this risk for Omicron patients was 0.78 (95% CI 0.62-0.97) and compared to ancestral lineages was 1.04 (95% CI 0.84-1.24). Among Omicron and Delta infections, patients with history of vaccination or prior COVID-19 had one-half the 14-day risk of severe disease or death (adjusted hazard ratio 0.46, IQR 0.34-0.62) but no significant outcome difference between Delta and Omicron infections.

**Conclusions:** Although the risk of severe disease or death for unvaccinated patients with Omicron was lower than Delta, it was similar to ancestral lineages. Severe outcomes were less common in vaccinated patients, but there was no difference between Delta and Omicron infections.

## INTRODUCTION

In December 2020, the novel SARS-CoV-2 lineage B.1.1.7, now known as Alpha variant was identified as the first Variant of Concern.^1^ Emerging SARS-CoV-2 variants, demonstrated most recently by Omicron continue to cause waves of infections and hospitalizations. Differences in the transmissibility and clinical severity of SARS-CoV-2 variants have the potential to greatly impact public health as large segments of the population simultaneously become infected and require hospitalization. By mid-January, 2022, daily hospitalizations in the United States for people with COVID-19 exceeded 150,000, higher than at any point during the pandemic.^2^

Omicron and Delta variants are more transmissible than earlier circulating SARS-CoV-2 lineages.^3,4^ Compared to previously circulating lineages, Alpha^5,6^ and Delta^7–10^ variants are associated with greater risk for hospitalization. Prior work has suggested that Delta variant infection is associated with worse outcomes for hospitalized patients. But these studies have been limited by sample size and clinical confounders.^7,8,11^ Recent studies report a decreased risk of hospitalization, severe illness, and death for Omicron compared to Delta variants, ^12–14^ but limited data are available to describe the clinical trajectory of patients once they become hospitalized.^12,15^

Changing demographics, comorbidities, vaccination status, and therapeutics confound interpretation of the impact of SARS-CoV-2 lineage on COVID-19 severity. As COVID-19 hospitalizations overwhelm health systems, there is a need to understand the clinical demands and outcomes of patients hospitalized with current variants such as Delta and Omicron.

Therefore, we used a clinical registry to observe the impact of SARS-CoV-2 variants on clinical trajectory and outcome.

## METHODS

### Ethical considerations and data availability

Research was conducted under Johns Hopkins institutional review board protocol IRB00300364 with a waiver of consent. SARS-CoV-2 genomes were uploaded to Global Initiative on Sharing All Influenza Data (GISAID).^16^

### Setting and data source

The data source used for this study is JH-CROWN, a registry of electronic medical record (EMR) data from Johns Hopkins Medicine (JHM), which includes five acute care hospitals in Maryland and the District of Columbia.^17^ The EMR includes SARS-CoV-2 test results and vaccination records from the Chesapeake Regional Information System for our Patients (CRISP)^18^ and other registries. Baseline comorbidities were determined from ICD-10 codes;^19^ conditions recorded as starting in prehospitalization encounters or with start dates <48 hours after hospitalization were considered.

### Cohort composition and outcome determination

Inclusion and exclusion criteria were designed to maximize inclusion of patients with COVID-19 severe enough to require inpatient care and minimize inclusion of patients with asymptomatic SARS-CoV-2 infections or COVID-19 of insufficient severity to independently warrant hospitalization. Inpatients with SARS-CoV-2 infection were identified based on positive JHM laboratory test results, record of positive SARS-CoV-2 test results from outside JHM recorded via CRISP, and identification by the local infection control team. Hospitalizations >14 days after diagnosis, >7 days prior to diagnosis, and subsequent hospitalizations after an index hospitalization were excluded. Symptomatic COVID-19 requiring inpatient care was defined by pulse oximetry recording of <95%, temperature ≥38°C, heart rate >110, respiratory rate >24, or use of supplemental oxygen within the first 24 hours of admission. Inpatients transferred from outside acute care facilities or admitted to surgical or psychiatric services were excluded.

Patients who received a dose of Ad26.COV2.S or a second dose of mRNA-1273 or BNT162b2 >2 weeks prior to hospitalization were classified as vaccinated; additional vaccine doses were not considered due to small sample size. Prior COVID-19 was defined by positive SARS-CoV-2 test >60 days prior to hospitalization. Unvaccinated patients with history of prior COVID-19 were grouped with vaccinated patients for stratified analyses based on reports showing similar protective effects.^20^

Severe COVID-19 was defined as respiratory support with high flow nasal cannula, noninvasive positive pressure ventilation, or mechanical ventilation.^21^ The primary outcome was severe disease or death within 14 days of hospital presentation, chosen because severe COVID-19 onset occurs within this timeframe for nearly all patients who develop severe disease.^17^ Patients who were discharged alive and without record of death were assumed to survive through day 14 without severe disease. Outcomes for patients who remained hospitalized were censored at the end of the observation period.

### SARS-CoV-2 whole genome sequencing (WGS)

WGS surveillance was performed on remnant positive SARS-CoV-2 clinical samples, chosen irrespective of hospitalization status with priority given to samples with cycle threshold values < 20. RNA was extracted using the Chemagic™ 360 system (Perkin Elmer) following the manufacturer’s protocol. WGS was performed on the Oxford Nanopore GridION using either the V3 primer ARTIC SARS-CoV-2 sequencing protocol or the NEBNext® ARTIC SARS-CoV-2 Companion Kit (VarSkip Short SARS-CoV-2 # E7660-L). Each GridION sequencing run pooled 94 samples plus a negative and positive control.^22^ Alignment and variant calling were performed with the artic-ncov2019 medaka protocol.^23^ Questionable mutations were visually inspected. Clades were determined using Nextclade,^24^ and lineages with Pangolin COVID-19 Lineage Assigner.^25^ Sequencing results with insufficient depth and coverage for clade and lineage assignment were excluded.

### Variant inference

The rapid spread of novel variants after local introduction defines time periods when a single viral clade dominates community transmission. Using WGS results from our health system, we determined the rolling two-week distribution of viral lineages classified as: Alpha variant, Delta variant, Omicron variant, ancestral lineages (emerging prior to Alpha) and other.^26^ When the rolling two-week average identified ≥95% of WGS results as a single variant, patients without WGS results who were diagnosed on that day were inferred to be infected with the dominant lineage.

### Statistical methods

Propensity score weighting was used to infer the causal effect of variant on outcomes. This approach infers the outcomes that might be observed if the entire cohort were infected with one variant (e.g. Delta) as compared to another (e.g. Omicron), or the average treatment effect of a particular variant. Only pre-infection baseline clinical features were considered as covariates in regression and propensity models, to avoid adjustment for intermediate variables in the causal pathway from the variant to outcomes that would bias estimates of the variants’ effects.

Propensity for a particular variant was assessed using gradient boosted logistic regression using the R twang package.^27^ Weighted regression models were fit using the survey package.^28^ Propensity models included age, sex, body mass index (BMI), Elixhauser score,^29^ and history of organ transplantation or immunocompromised status. Log binomial regression assessed the average treatment effect on the Risk Difference and Relative Risk scales. Models estimated remdesivir use, steroid use, and hypoxia at presentation, reported by the ratio of oxygen saturation by pulse oximetry to the fraction of inspired oxygen (SpO_2_/FiO_2_). The probability of surviving and remaining free of severe disease was assessed using the Kaplan Meier estimator. Convergence, positivity, and balance were assessed in propensity score analyses.

Sensitivity analysis restricted to patients with confirmed variants was performed using age, sex, and Elixhauser score to determine propensity scores. The association of vaccination or prior COVID-19 with severe disease was limited to patients with Omicron or Delta infections and assessed using a Cox proportional hazard model adjusted for baseline clinical features, variant, and the interaction of variant and history of vaccination or prior COVID-19.

## RESULTS

### Patient population

From September 1, 2020, to January 20, 2022, 7,038 adults were hospitalized within 2 weeks of COVID-19 diagnosis; 297 patients transferred from other acute care facilities and 203 admitted to surgical or psychiatric services who were excluded (**Supplementary Figure 1**). Among 6,538 remaining patients, 5,706 (87%) had vital sign abnormalities of tachycardia (n =2,079, 36%), tachypnea (n = 2,866, 50%), fever (n = 2,055, 36%), hypoxia (n = 4,543, 80%), or requirement for supplemental oxygen (n=3,802, 67%) and were considered for inclusion.

### WGS results and variant inference

For 285 of 506 (56.3%) days of study, a single variant group accounted for ≥95% of the rolling two-week average (**Figure 1 and Supplementary Figure 2**) including ancestral lineages (139 days), Delta (124 days), and Omicron (22 days). Among 5,706 patients meeting clinical inclusion criteria, WGS results were available for 1,111 (19%) inpatients; variant was inferred for an additional 3,062 (54%) inpatients. Confirmed or inferred variant data was available for 3,365 of 4,667 (72%) unvaccinated and 808 of 1030 (78%) vaccinated or previously infected patients. Among 575 included patients with available WGS results during a period when a single lineage was dominant, inferred variant matched the WGS result for 553 (96%) patients. During the period of Omicron dominance, 111 of 128 (87%) patients with available WGS results were Omicron including 29 of 36 (81%) patients with severe disease.

**Figure 1.**
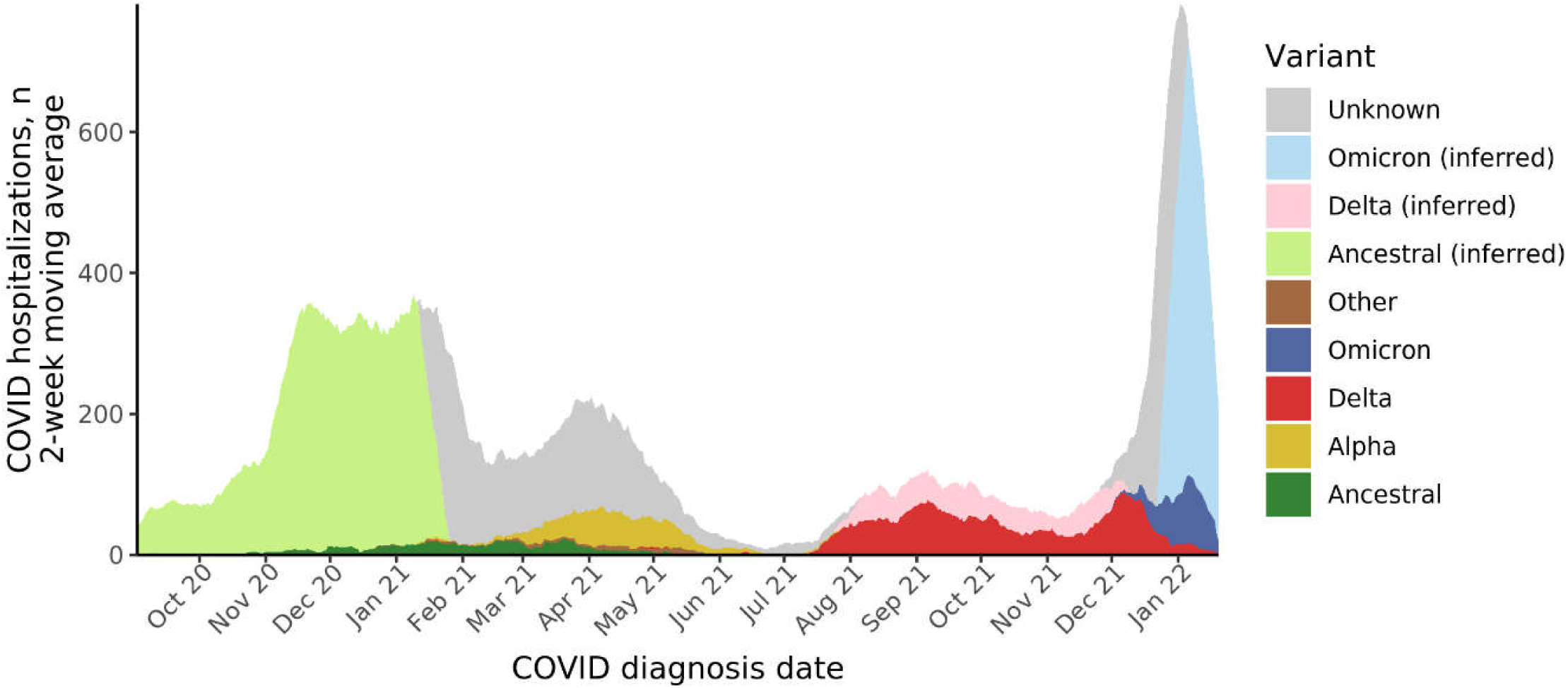
SARS-CoV-2 Variants Among Hospitalized Patients, Two-Week Moving Average by Date of COVID-19 Diagnosis and Variant COVID-19-related hospitalizations with available confirmed SARS-CoV-2 variant results determined by whole-genome sequencing (WGS) are shaded in darker colors. WGS surveillance of positive SARS-CoV-2 samples within JHM was used to determine a community prevalence of SARS-CoV-2 variants (**Supplemental Figure 1**). During time periods when the two-week moving distribution of variants reached 95%, variant was inferred for hospitalized patients who did not have WGS results. During periods when no single variant reached 95%, no inference of variant for patient without WGS was attempted.

**Figure 2:**
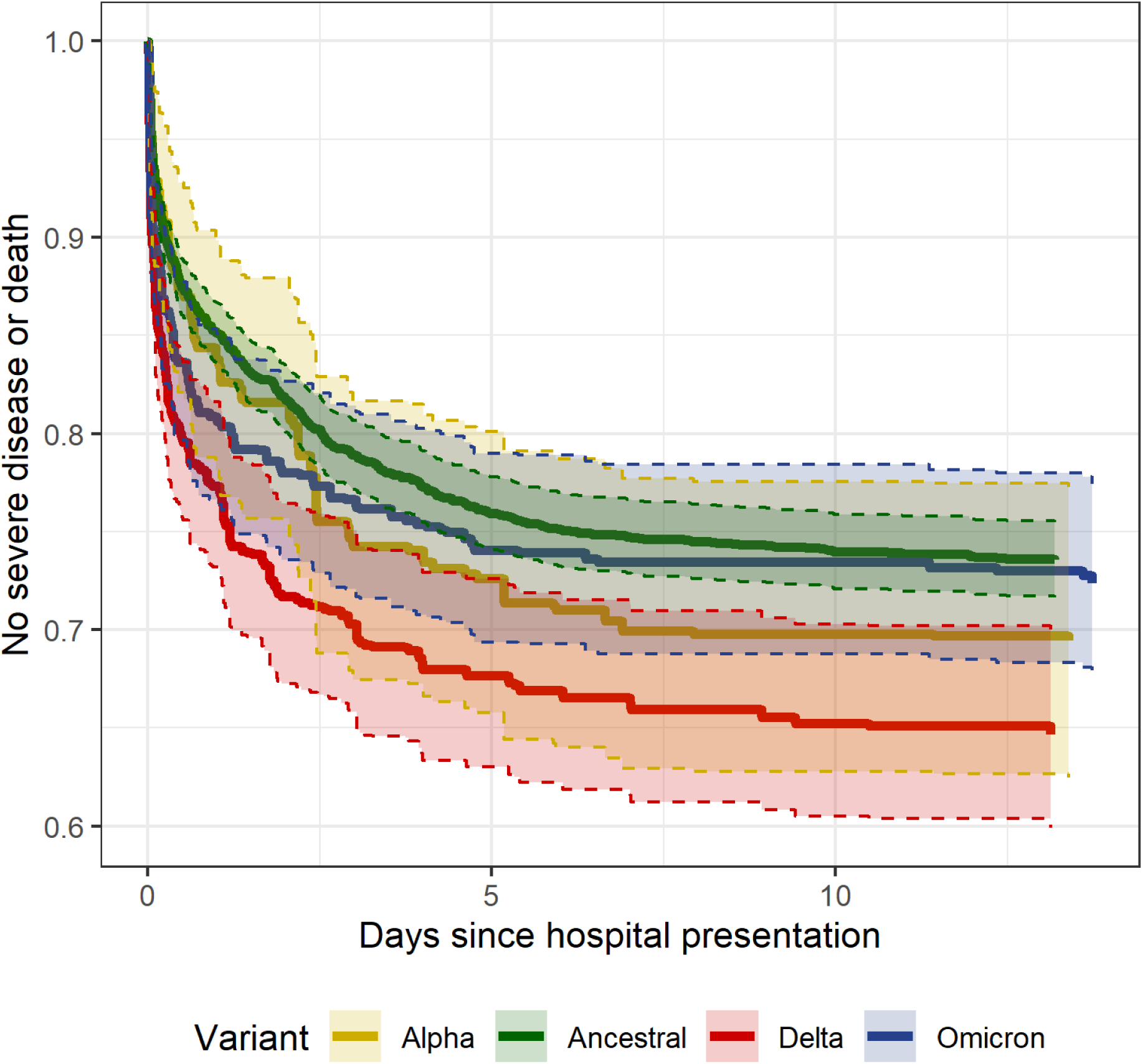
Inverse probability weighted risk of severe disease or death by days since hospital presentation and variant among 3,198 unvaccinated patients hospitalized with COVID-19. Among 3365 unvaccinated patients with available inferred variant data, 34 with other variants and 133 with missing BMI were excluded from the model.

### Clinical and demographic features

Among 3,365 unvaccinated patients, median age was 62 years (interquartile range [IQR] 49-75) (**Table 1**), 1,691 (50%) were male, and median BMI was 28.5 (IQR 24.4-34). Vaccinated patients were older, with median age 69 years (IQR 58-79, p < 0.001, **Supplementary Table 1**). Patients with history of vaccination or prior COVID-19 had more comorbidities than unvaccinated patients (median Elixhauser score of 6; IQR 4-9, compared to 5; IQR 3-7). Among the unvaccinated, patients infected with ancestral lineages were older than other groups (median age 65, IQR 52-78, compared to 58, IQR 43-70). The proportion of patients who were Black was higher in Alpha (63%), Delta (42%), and Omicron (46%) variant groups than in ancestral lineages (31%).

**Table 1.** Demographics and clinical characteristics by variant among 3,365 unvaccinated patients hospitalized with COVID-19

|  | All<br>N=3365 | Ancestral<br>N=2116 | Alpha<br>N=214 | Delta<br>N=568 | Omicron<br>N=433 | Other<br>N=34 |
| --- | --- | --- | --- | --- | --- | --- |
| Age, y | 62.0 (49.0-75.0) | 65.0 (52.0-78.0) | 59.0 (47.2-69.0) | 55.0 (41.0-67.0) | 62.0 (45.0-73.0) | 61.0 (50.2-66.2) |
| Sex: |  |  |  |  |  |  |
| Female | 1674 (49.7%) | 1042 (49.2%) | 110 (51.4%) | 280 (49.3%) | 224 (51.7%) | 18 (52.9%) |
| Male | 1691 (50.3%) | 1074 (50.8%) | 104 (48.6%) | 288 (50.7%) | 209 (48.3%) | 16 (47.1%) |
| White blood cell count | 6.7 (5.0-9.1) | 6.6 (5.0-9.1) | 6.2 (4.8-8.3) | 6.2 (4.7-8.7) | 7.7 (5.4-10.4) | 6.0 (3.9-6.8) |
| Body mass index, kg/m<sup>2</sup> | 28.5 (24.4-34.0) | 28.5 (24.5-33.8) | 29.5 (24.9-35.3) | 28.9 (24.4-35.0) | 27.4 (23.3-33.3) | 31.8 (26.4-37.4) |
| Race/ethnicity: |  |  |  |  |  |  |
| Asian | 163 (4.8%) | 128 (6.0%) | 8 (3.7%) | 17 (3.0%) | 9 (2.1%) | 1 (2.9%) |
| Black | 1248 (37.1%) | 661 (31.2%) | 135 (63.1%) | 237 (41.7%) | 201 (46.4%) | 14 (41.2%) |
| Hispanic | 388 (11.5%) | 289 (13.7%) | 7 (3.3%) | 48 (8.5%) | 43 (9.9%) | 1 (2.9%) |
| Non-Hispanic white | 1406 (41.8%) | 938 (44.3%) | 56 (26.2%) | 239 (42.1%) | 158 (36.5%) | 15 (44.1%) |
| Other or unknown | 160 (4.8%) | 100 (4.7%) | 8 (3.7%) | 27 (4.8%) | 22 (5.1%) | 3 (8.8%) |
| Nursing home admission | 221 (6.6%) | 196 (9.3%) | 3 (1.4%) | 9 (1.6%) | 13 (3.0%) | 0 (0.0%) |
| Elixhauser score | 5.0 (3.0-7.0) | 5.0 (3.0-7.0) | 5.0 (3.0-8.0) | 4.0 (2.0-6.0) | 4.0 (3.0-7.0) | 4.0 (3.0-8.5) |
| Immunocompromised or transplant | 288 (8.6%) | 173 (8.2%) | 38 (17.8%) | 45 (7.9%) | 30 (6.9%) | 2 (5.9%) |
| Diabetes | 1334 (39.6%) | 874 (41.3%) | 92 (43.0%) | 190 (33.5%) | 165 (38.1%) | 13 (38.2%) |
| Cardiovascular disease | 1594 (47.4%) | 1049 (49.6%) | 108 (50.5%) | 211 (37.1%) | 213 (49.2%) | 13 (38.2%) |
| Chronic pulmonary disease | 1094 (32.5%) | 710 (33.6%) | 78 (36.4%) | 167 (29.4%) | 127 (29.3%) | 12 (35.3%) |
| Malignancy | 948 (28.2%) | 643 (30.4%) | 73 (34.1%) | 116 (20.4%) | 104 (24.1%) | 12 (35.3%) |
| Pregnant | 25 (0.7%) | 12 (0.6%) | 2 (0.9%) | 7 (1.2%) | 4 (0.9%) | 0 (0.0%) |
| SpO<sub>2</sub>/FiO<sub>2</sub> ratio | 435.0 (275.0-459.0) | 430.0 (281.0-454.0) | 440.0 (293.8-459.0) | 430.0 (261.0-454.0) | 444.0 (277.0-464.0) | 442.0 (336.0-459.0) |
| Pulse | 98.0 (85.0-112.0) | 97.0 (84.0-111.0) | 101.5 (90.0-113.0) | 103.0 (88.0-115.0) | 101.0 (86.0-114.0) | 93.5 (85.0-107.2) |
| Respiratory rate, breaths/min | 20.0 (18.0-26.0) | 21.0 (18.0-27.0) | 20.0 (18.0-25.0) | 20.0 (18.0-26.0) | 20.0 (18.0-26.0) | 21.0 (18.0-27.5) |
| Temperature, °C | 37.2 (36.8-37.9) | 37.2 (36.8-37.9) | 37.3 (36.8-38.2) | 37.1 (36.7-37.9) | 36.9 (36.5-37.5) | 37.5 (37.0-38.2) |
| GFR, mL/min per 1.73 m<sup>2</sup> | 70.0 (45.0-94.0) | 68.0 (44.0-91.0) | 63.5 (36.2-92.0) | 82.0 (53.0-99.0) | 70.0 (46.0-99.0) | 83.0 (63.5-90.0) |
| Albumin level, g/L | 3.7 (3.3-4.1) | 3.7 (3.4-4.1) | 3.8 (3.3-4.2) | 3.8 (3.2-4.1) | 3.7 (3.1-4.1) | 3.8 (3.4-4.2) |
| ALT level, U/L | 27.0 (18.0-45.2) | 27.0 (17.0-44.0) | 25.0 (17.0-41.0) | 30.0 (20.0-52.0) | 27.0 (17.0-47.0) | 24.5 (16.2-42.8) |
| Total bilirubin level, mg/dL | 0.5 (0.3-0.7) | 0.5 (0.3-0.7) | 0.5 (0.4-0.7) | 0.5 (0.3-0.7) | 0.5 (0.3-0.8) | 0.4 (0.3-0.7) |
| CRP level, mg/L | 6.3 (2.6-11.7) | 6.2 (2.5-11.4) | 5.8 (3.0-11.1) | 7.4 (3.3-12.2) | 6.0 (2.0-11.5) | 3.5 (1.3-10.1) |
| IL-6 level, pg/mL | 28.5 (11.4-63.9) | 27.0 (11.3-59.2) | 17.8 (10.7-80.1) | 36.8 (12.9-94.4) | 52.9 (27.8-89.9) | 36.5 (26.4-11026.9) |

Vital sign and labs are the first observation within 36 hours of hospital admission. Unvaccinated persons with a documented history of COVID-19 infection >60 days prior to hospitalization were excluded. SpO<sub>2</sub>/FiO<sub>2</sub> ratio, ratio of oxygen saturation by pulse oximetry to the fraction of inspired oxygen; GFR, glomerular filtration rate; ALT, alanine aminotransferase; CRP, C-reactive protein; IL-6, interleukin-6

**Table 2.** Severe disease or death by day 14 of hospitalization among hospitalized unvaccinated patients

|  | All patients<br>N=3,365 | No severe disease<br>or death<br>N=2,415 | Severe disease or<br>death<br>N=950 | p |
| --- | --- | --- | --- | --- |
| Age, y | 62.0 (49.0-75.0) | 61.0 (47.0-74.0) | 65.0 (55.0-77.0) | <0.001 |
| Sex: |  |  |  | <0.001 |
| Female | 1674 (49.7%) | 1255 (52.0%) | 419 (44.1%) |  |
| Male | 1691 (50.3%) | 1160 (48.0%) | 531 (55.9%) |  |
| White blood cell count | 6.7 (5.0-9.1) | 6.4 (4.8-8.5) | 7.6 (5.5-10.7) | <0.001 |
| Body mass index, kg/m<sup>2</sup> | 28.5 (24.4-34.0) | 28.2 (24.2-33.5) | 29.5 (25.0-35.8) | <0.001 |
| Race/ethnicity: |  |  |  | <0.001 |
| Asian | 163 (4.8%) | 126 (5.2%) | 37 (3.9%) |  |
| Black | 1248 (37.1%) | 941 (39.0%) | 307 (32.3%) |  |
| Hispanic | 388 (11.5%) | 290 (12.0%) | 98 (10.3%) |  |
| Non-Hispanic white | 1406 (41.8%) | 944 (39.1%) | 462 (48.6%) |  |
| Other or unknown | 160 (4.8%) | 114 (4.7%) | 46 (4.8%) |  |
| Nursing home admission | 221 (6.6%) | 127 (5.3%) | 94 (9.9%) | <0.001 |
| Elixhauser score | 5.0 (3.0-7.0) | 4.0 (2.0-7.0) | 6.0 (4.0-8.0) | <0.001 |
| Immunocompromised or transplant | 288 (8.6%) | 198 (8.2%) | 90 (9.5%) | 0.262 |
| Diabetes | 1334 (39.6%) | 892 (36.9%) | 442 (46.5%) | <0.001 |
| Cardiovascular disease | 1594 (47.4%) | 1017 (42.1%) | 577 (60.7%) | <0.001 |
| Chronic pulmonary disease | 1094 (32.5%) | 723 (29.9%) | 371 (39.1%) | <0.001 |
| Malignancy | 948 (28.2%) | 686 (28.5%) | 262 (27.6%) | 0.637 |
| Pregnant | 25 (0.7%) | 21 (0.9%) | 4 (0.4%) | 0.254 |
| SpO<sub>2</sub>/FiO<sub>2</sub> ratio | 435.0 (275.0-459.0) | 449.0 (342.0-459.0) | 235.0 (107.0-346.0) | <0.001 |
| Pulse | 98.0 (85.0-112.0) | 97.0 (84.0-111.0) | 102.0 (88.0-116.0) | <0.001 |
| Respiratory rate, breaths/min | 20.0 (18.0-26.0) | 20.0 (18.0-24.0) | 25.5 (20.0-33.0) | <0.001 |
| Temperature, C | 37.2 (36.8-37.9) | 37.1 (36.7-37.9) | 37.2 (36.8-38.1) | 0.012 |
| GFR, mL/min per 1.73 m<sup>2</sup> | 70.0 (45.0-94.0) | 75.0 (49.0-97.0) | 58.0 (36.0-84.0) | <0.001 |
| Albumin level, g/L | 3.7 (3.3-4.1) | 3.8 (3.4-4.2) | 3.5 (3.0-3.9) | <0.001 |
| ALT level, U/L | 27.0 (18.0-45.2) | 26.0 (17.0-43.0) | 30.0 (20.0-50.0) | <0.001 |
| Total bilirubin level, mg/dL | 0.5 (0.3-0.7) | 0.5 (0.3-0.7) | 0.5 (0.4-0.8) | <0.001 |
| CRP level, mg/L | 6.3 (2.6-11.7) | 5.0 (1.8-9.4) | 9.9 (5.4-16.2) | <0.001 |
| IL-6 level, pg/mL | 28.5 (11.4-63.9) | 22.1 (9.4-46.5) | 46.8 (20.0-109.5) | <0.001 |
| Variant, inferred or confirmed |  |  |  | 0.171 |
| Ancestral | 2116 (62.9%) | 1537 (63.6%) | 579 (60.9%) |  |
| Alpha | 214 (6.4%) | 148 (6.1%) | 66 (6.9%) |  |
| Delta | 568 (16.9%) | 386 (16.0%) | 182 (19.2%) |  |
| Omicron | 433 (12.9%) | 319 (13.2%) | 114 (12.0%) |  |
| Other | 34 (1.0%) | 25 (1.0%) | 9 (0.9%) |  |
| Variant, confirmed only |  |  |  | 0.695 |
| Ancestral | 152 (18.7%) | 110 (19.7%) | 42 (16.5%) |  |
| Alpha | 214 (26.3%) | 148 (26.5%) | 66 (25.9%) |  |
| Delta | 351 (43.1%) | 233 (41.7%) | 118 (46.3%) |  |
| Omicron | 63 (7.7%) | 43 (7.7%) | 20 (7.8%) |  |
| Other | 34 (4.2%) | 25 (4.5%) | 9 (3.5%) |  |

SpO<sub>2</sub>/FiO<sub>2</sub> ratio, ratio of oxygen saturation by pulse oximetry to the fraction of inspired oxygen; GFR, glomerular filtration rate; ALT, alanine aminotransferase; CRP, C-reactive protein; IL-6, interleukin-6

Presenting SpO_2_/FiO_2_ was higher for patients with Omicron infections (median 444, IQR 277-464) compared to patients with Delta (median 430, IQR 261-454) and ancestral lineage infections (median 430, IQR 281-454). Among 2,563 (76%) unvaccinated patients with C-reactive protein (CRP) recorded <36 hours after admission, patients with Omicron infections had lower (5.95, IQR 2.02-11.5) and Delta patients had higher (7.38, IQR 3.3-12.2) median CRP levels, compared to patients with ancestral lineage infections (6.24, IQR 2.5-11.45).

### Inpatient outcomes, unvaccinated

Inpatient outcomes were observed through January 29, 2022. On that day, 5 unvaccinated patients remained hospitalized for < 14 days and not severely ill. Among unvaccinated patients, 950 (28%) developed severe disease or death within 14 days of hospitalization (median time to event 0.47 days, IQR 0.07-2.41) including 579 (27%) ancestral, 66 (31%) Alpha, 182 (32%) Delta, 114 (26%) Omicron, and 9 (26%) other variant group infections. By 14-days of follow-up, 125 (4%) patients remained at risk for incident severe disease. There were 525 (16%) unvaccinated patients who required mechanical ventilation or died by hospital day 21 (median time to event 2.82 days, IQR 0.52-7.92, **Supplementary Figure 3**). By day 30, there were 194 (9%) ancestral, 11 (5%) Alpha, 46 (8%) Delta, 27 (6%) Omicron, and 3 (9%) other patients who died (median time to death 10.32 days, IQR 5.31-17.63).

Average treatment effect models estimated a 14-day probability of severe disease or death by variant group of 0.26 for ancestral, 0.31 for Alpha, 0.35 for Delta, and 0.28 for Omicron infections (**Table 3**, model balance metrics shown in **Supplementary Table 2** and **Figure 4**). The relative risk of developing severe disease or death within 14 days for Delta variant compared to ancestral lineages was 1.34 (95% confidence interval [CI] 1.13-1.55). Compared to Delta variant, the 14-day relative risk of severe disease or death for Omicron was 0.78 (95% CI 0.62-0.97) and compared to ancestral lineages was 1.04 (0.84-1.24). Compared to ancestral lineages, the weighted mean presenting SpO_2_/FiO_2_ for Delta variant was lower by 22.07 (95% CI −36.37 to −7.77). Patients with Delta infections were more likely to receive remdesivir (relative risk 1.11, 95% CI 1.02-1.22) and steroids (relative risk 1.06, 95% CI 1-1.13) and Omicron patients less likely to receive steroids (relative risk 0.9, 95% CI 0.83-0.97) than patients with ancestral lineage infections. A sensitivity analysis of 2,744 unvaccinated patients that excluded inference of Alpha, Delta, and Omicron variants showed similar estimates for the relative risk of severe disease or death, but with wider confidence intervals (**Supplementary Table 3** and **Figure 5**).

**Table 3:** Clinical outcomes and features by variant in an average treatment effect model adjusted for risk factors for severe COVID-19.

|  | **Ancestral** | **Alpha** | **Delta** | **Omicron** |
| --- | --- | --- | --- | --- |
| Mean presenting SpO<sub>2</sub>/FiO<sub>2</sub> | 364.6<br>(359.53, 369.67) | 363.25<br>(344.63, 381.87), | 342.53<br>(329.16, 355.9), | 361.38<br>(347.78, 374.98), |
| Mean Difference | Ref | -1.35<br>(-20.65, 17.95) | -22.07<br>(-36.37, -7.77) | -3.22<br>(-17.73, 11.29) |
| 14-day risk of severe disease or death |  |  |  |  |
| Estimate | 0.26 (0.25, 0.28) | 0.31 (0.23, 0.38) | 0.35 (0.3, 0.4) | 0.28 (0.23, 0.32) |
| Relative risk, Ancestral as reference | Ref | 1.16 (0.86, 1.45) | 1.34 (1.13, 1.55) | 1.04 (0.84, 1.24) |
| Relative risk, Delta as reference | 0.75 (0.64-0.87) | 0.87 (0.65-1.15) | Ref | 0.78 (0.62-0.97) |
| Risk difference, Ancestral as reference | Ref | 0.04 (-0.04, 0.12) | 0.09 (0.04, 0.14) | 0.01 (-0.04, 0.06) |
| Treatment with remdesivir |  |  |  |  |
| Estimate | 0.53 (0.51, 0.55) | 0.6 (0.52, 0.68) | 0.59 (0.54, 0.64) | 0.53 (0.48, 0.59) |
| Treatment with dexamethasone |  |  |  |  |
| Estimate | 0.72 (0.7, 0.74) | 0.77 (0.7, 0.83) | 0.76 (0.72, 0.8) | 0.64 (0.59, 0.69) |

The average treatment effect model uses inverse probability weighting to estimate clinical features and outcomes if the entire cohort was composed of a single variant. Weighting was performed using risk factors for severe COVID-19 including age, sex, body mass index, Elixhauser comorbidity score, and presence of immunosuppression or history of transplantation.

### Inpatient outcomes, patients with history of vaccination or prior COVID-19

The 808 patients with history of vaccination or prior COVID-19 included 291 (34%) with Delta and 500 (62%) with Omicron infections; only 17 vaccinated or previously infected patients with Alpha, Ancestral, or other lineages were hospitalized. There were 67 (23%) vaccinated Delta and 109 (22%) vaccinated Omicron patients who developed severe disease or death within 14 days (**Supplementary Table 4**) with median time to event of 0.52 (IQR 0.09-3.15) days. By 14 days of follow up, there remained 43 (5%) vaccinated persons at risk for incident severe disease or death.

Among patients with Omicron and Delta infections, inpatients who were vaccinated or had history of COVID-19 had half the 14-day risk of severe disease or death compared to unvaccinated inpatients (adjusted hazard ratio 0.46, IQR 0.34-0.62, **Supplementary Table 5**). The adjusted 14-day risk of severe disease or death was similar between patients with Delta variant and Omicron variant infections (**Supplementary Figure 6**). There were 33 (11%) vaccinated Delta and 53 (11%) vaccinated Omicron patients who required mechanical ventilation or died within the first 14 days of hospitalization (median time to event 2.89 days, IQR 0.22-7.72).

## DISCUSSION

Leveraging on-site SARS-CoV-2 genomic surveillance and real-time linkage to EMR, we report clinically relevant differences in the risk for developing severe disease among patients hospitalized with symptomatic COVID-19 by SARS-CoV-2 variant. Compared to previously circulating lineages, unvaccinated patients infected with Delta variant were more likely to develop severe illness or death when adjusted for known risk factors for severe COVID-19.

Although unvaccinated patients hospitalized with Omicron variant had a lower risk of developing severe disease or death compared to those with Delta variant, the risk of severe disease or death was similar compared to SARS-CoV-2 lineages that circulated earlier in the pandemic. Among vaccinated patients, there was no difference in the risk of developing severe illness between Delta and Omicron variants once patients were hospitalized.

The finding that unvaccinated individuals currently hospitalized with COVID-19 amidst Omicron-dominant community transmission do not have significantly decreased risk of developing severe disease compared to cases prior to emergence of variants of concern undercuts public perception that Omicron is a mild disease. Reports from South Africa, Scotland, and Southern California using inferred variant data from S gene target failure on PCR have shown that compared to patients with Delta infections, outpatients infected with Omicron have a 0.2-0.3-fold risk of requiring hospitalization.^12–14^ Once hospitalized, patients with Omicron in the South African cohort had a 0.3-fold risk of developing severe disease compared to patients with Delta, but this study included any oxygen therapy as a criteria for severe disease.^14^ In southern California, there was a 0.26-fold risk of intensive care unit admission among Omicron compared to Delta patients. Comparison of disease severity between Omicron and Delta alone fails to account for the increased severity of Delta variant compared to prior lineages which were responsible for millions of deaths. Using stricter criteria for cohort entry designed to exclude asymptomatic SARS-CoV-2 infection, we found a 0.78-fold risk of developing severe disease for patients with Omicron compared to Delta, but no difference compared to patients with ancestral lineages.

Patients who were vaccinated or previously had COVID-19 were only half as likely as unvaccinated patients to develop severe disease in the era of Delta and Omicron transmission, and there was no difference in this risk by variant. We found that the vaccinated patients who did require hospitalization had more advanced age and comorbidities than unvaccinated inpatients. Vaccines remain largely effective in preventing COVID-19 related hospitalization and death, even after the emergence of Delta and Omicron variants.^30,31^ Most importantly, population-based surveillance shows a striking 16-fold greater risk of hospitalization for unvaccinated individuals compared to the vaccinated.^32^

Comparison of the risk of developing severe disease between variants is critical as the evidence base for understanding disease severity, therapeutic efficacy, and health system burden is primarily based on observations recorded before the emergence of Delta variant. The perception that young and healthy persons who require hospitalization for COVID-19 are unlikely to require ICU-level care formed during the first year of the pandemic underestimated their risk after Delta variant emergence. As it is not feasible to repeat the study of every COVID-19 therapeutic each time a new variant emerges, data comparing the risk of developing severe disease by variant is necessary for clinicians to regauge the risks and benefits of COVID-19 therapeutics.

Study limitations include reliance on community prevalence to infer SARS-CoV-2 lineage for most patients. However, surveillance was performed locally with 96% concordance of inferred to confirmed variant. Model estimates from sensitivity analyses limited to confirmed variants were similar to primary analyses. Co-circulation of the comparatively severe Delta variant at low levels during Omicron-dominant transmission may overrepresent Delta among hospitalized patients, especially the severely ill. Although inferred-confirmed variant concordance decreased to 87% during Omicron-dominant transmission, this misclassification biases against finding a severity difference between Omicron and Delta. Differences in patient characteristics and care patterns may limit the generalizability of estimates of severe disease risk. Reliance on vital sign abnormalities likely includes patients whose hospitalization may be due to exacerbation of chronic medical conditions temporally related to a positive SARS-CoV-2 test and misses others with more subtle COVID-19 symptoms.

Study strengths include strict, objective inclusion criteria for symptomatic COVID-19, detailed clinical information from the EMR, extensive WGS results, and robust causal inference methods. Our findings show that for vaccinated patients who are hospitalized with COVID-19, their risk of severe disease or death is low–reduced by half compared to unvaccinated patients - and equally low for both Omicron and Delta infections. Despite the comparatively lower risk of hospitalization for Omicron infections, for unvaccinated patients who require hospitalization, there remains a risk of severe disease nearly equivalent to pre-Delta waves of COVID-19 infections.

## Data Availability

SARS-CoV-2 genomes were uploaded to Global Initiative on Sharing All Influenza Data (GISAID).

## ACKNOWLEDGMENTS

We acknowledge the Johns Hopkins Precision Medicine Analytics Platform (PMAP) and Core for Clinical Research Data Acquisition (CCDA) for preparing the EMR data for analysis. This study was only possible with the efforts of the Johns Hopkins Clinical Microbiology Laboratory and clinicians and staff caring for patients with COVID-19.

## Supplementary Material

**Supplementary Figure 1.**
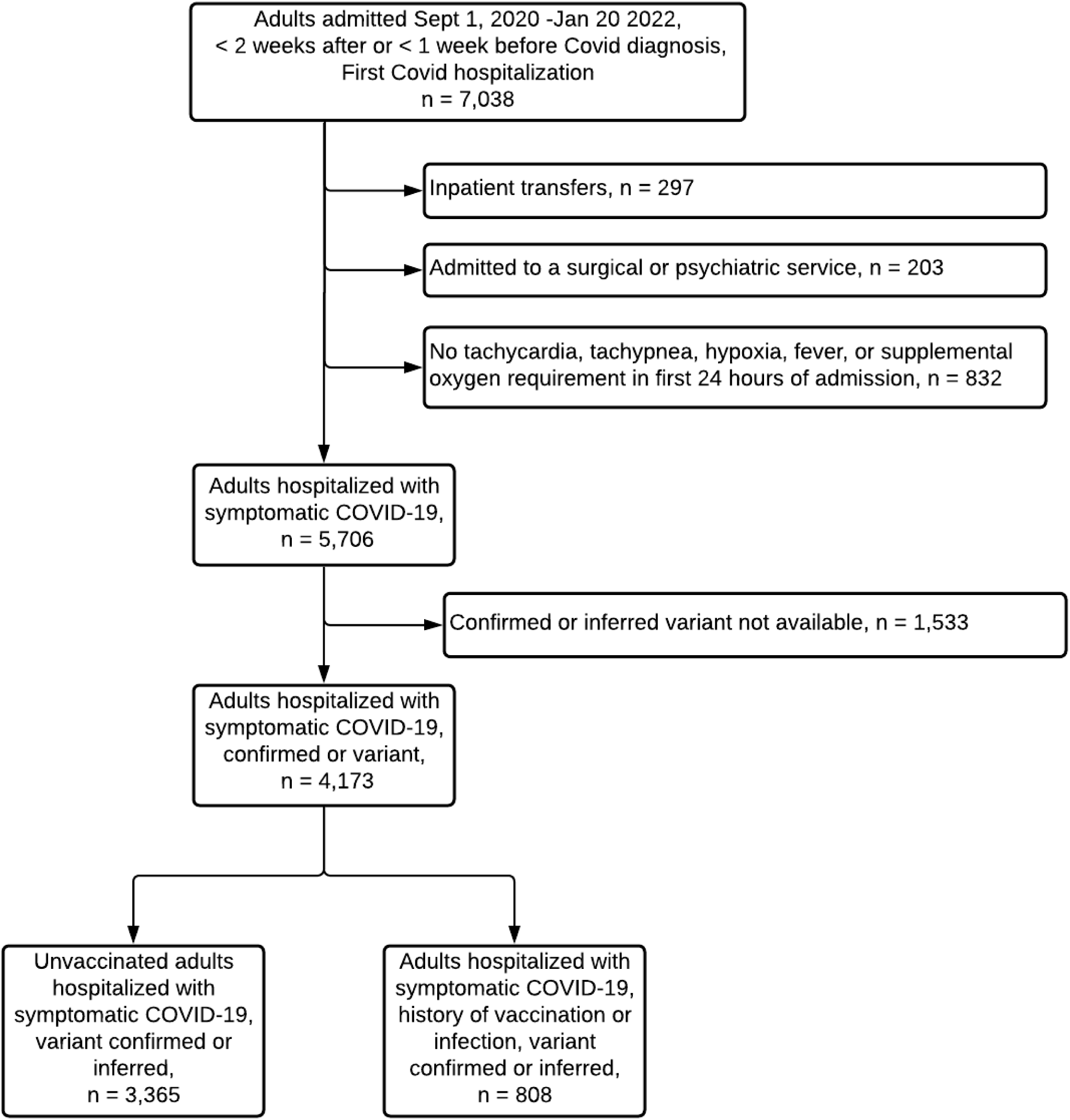
Cohort composition.

**Supplementary Figure 2.**
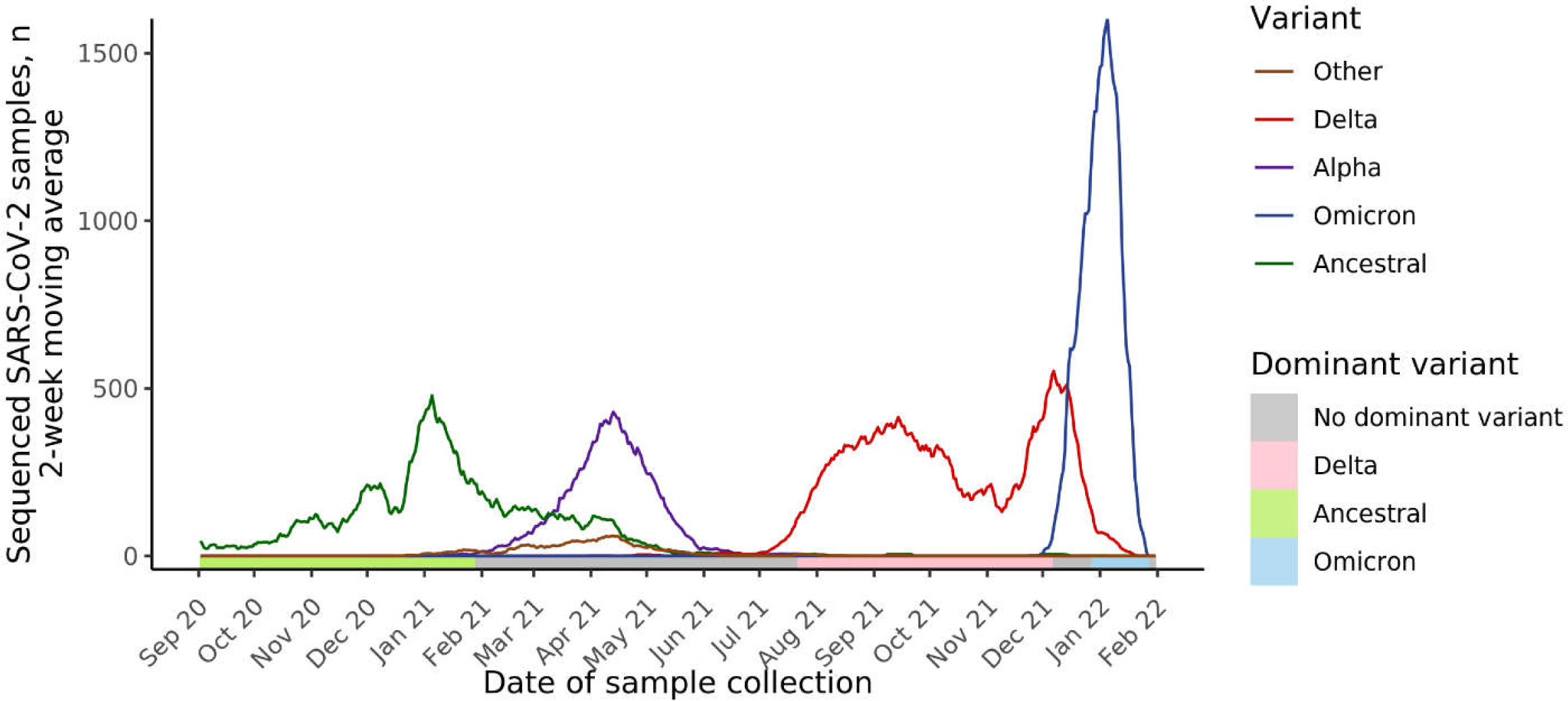
Distribution of variants over time for all sequenced samples. Two-week moving average of SARS-CoV-2 samples by date of sample collection and variant.

**Supplementary Table 1.** Demographics and clinical characteristics of hospitalized patients by variant among 808 patients hospitalized with COVID-19 with history of vaccination or prior COVID

|  | All patients<br>N=808 | Ancestral<br>N=5 | Alpha<br>N=9 | Delta<br>N=291 | Omicron<br>N=500 | Other<br>N=3 |
| --- | --- | --- | --- | --- | --- | --- |
| Vaccinated | 801 (99.1%) | 2 (40.0%) | 8 (88.9%) | 290 (99.7%) | 499 (99.8%) | 2 (66.7%) |
| Prior COVID-19 | 8 (1.0%) | 3 (60.0%) | 1 (11.1%) | 2 (0.7%) | 1 (0.2%) | 1 (33.3%) |
| Age, y | 69.0 (58.0-79.0) | 58.0 (53.0-66.0) | 59.0 (55.0-66.0) | 68.0 (59.0-80.0) | 69.0 (57.0-79.0) | 74.0 (65.5-82.5) |
| Sex: |  |  |  |  |  |  |
| Female | 382 (47.3%) | 3 (60.0%) | 6 (66.7%) | 141 (48.5%) | 230 (46.0%) | 2 (66.7%) |
| Male | 426 (52.7%) | 2 (40.0%) | 3 (33.3%) | 150 (51.5%) | 270 (54.0%) | 1 (33.3%) |
| White blood cell count | 7.4 (5.3-10.3) | 8.3 (6.4-8.8) | 4.3 (2.6-9.1) | 7.0 (5.3-9.3) | 7.7 (5.4-10.9) | 4.2 (3.9-4.7) |
| Body mass index, kg/m<sup>2</sup> | 27.9 (23.9-33.3) | 33.7 (31.4-41.3) | 27.3 (19.7-37.7) | 28.9 (24.4-34.3) | 27.1 (23.6-32.7) | 32.0 (29.2-33.5) |
| Race/ethnicity: |  |  |  |  |  |  |
| Asian | 31 (3.8%) | 1 (20.0%) | 0 (0.0%) | 17 (5.8%) | 13 (2.6%) | 0 (0.0%) |
| Black | 281 (34.8%) | 2 (40.0%) | 6 (66.7%) | 79 (27.1%) | 193 (38.6%) | 1 (33.3%) |
| Hispanic | 46 (5.7%) | 1 (20.0%) | 1 (11.1%) | 11 (3.8%) | 33 (6.6%) | 0 (0.0%) |
| Non-Hispanic white | 422 (52.2%) | 1 (20.0%) | 2 (22.2%) | 173 (59.5%) | 244 (48.8%) | 2 (66.7%) |
| Other or unknown | 28 (3.5%) | 0 (0.0%) | 0 (0.0%) | 11 (3.8%) | 17 (3.4%) | 0 (0.0%) |
| Nursing home admission | 41 (5.1%) | 0 (0.0%) | 0 (0.0%) | 11 (3.8%) | 30 (6.0%) | 0 (0.0%) |
| Elixhauser score | 6.0 (4.0-9.0) | 8.0 (3.0-9.0) | 7.0 (4.0-10.0) | 6.0 (4.0-9.0) | 6.0 (4.0-9.0) | 3.0 (2.0-6.5) |
| Immunocompromised or transplant | 178 (22.0%) | 1 (20.0%) | 1 (11.1%) | 73 (25.1%) | 102 (20.4%) | 1 (33.3%) |
| Diabetes | 394 (48.8%) | 3 (60.0%) | 4 (44.4%) | 145 (49.8%) | 242 (48.4%) | 0 (0.0%) |
| Cardiovascular disease | 521 (64.5%) | 3 (60.0%) | 5 (55.6%) | 180 (61.9%) | 331 (66.2%) | 2 (66.7%) |
| Chronic pulmonary disease | 334 (41.3%) | 3 (60.0%) | 4 (44.4%) | 116 (39.9%) | 210 (42.0%) | 1 (33.3%) |
| Malignancy | 337 (41.9%) | 1 (20.0%) | 4 (44.4%) | 139 (47.9%) | 192 (38.6%) | 1 (33.3%) |
| SpO<sub>2</sub>/FiO<sub>2</sub> ratio | 444.0 (327.5-459.0) | 449.0 (357.0-459.0) | 435.0 (213.0-444.0) | 444.0 (339.0-459.0) | 449.0 (308.2-464.0) | 357.0 (353.0-396.0) |
| Pulse | 96.0 (82.0-111.0) | 87.0 (82.0-99.0) | 101.0 (99.0-111.0) | 92.0 (80.5-109.5) | 98.0 (83.0-112.0) | 102.0 (82.5-122.5) |
| Respiratory rate, breaths/min | 20.0 (18.0-24.0) | 18.0 (17.0-22.0) | 24.0 (19.0-39.0) | 20.0 (18.0-24.0) | 20.0 (18.0-24.0) | 18.0 (17.0-21.0) |
| Temperature, C | 36.9 (36.6-37.5) | 37.0 (36.8-37.0) | 36.9 (36.7-38.6) | 37.0 (36.7-37.7) | 36.9 (36.6-37.3) | 37.2 (36.5-37.9) |
| GFR, mL/min per 1.73 m<sup>2</sup> | 62.0 (37.0-87.0) | 35.0 (6.0-89.0) | 58.0 (49.0-102.0) | 62.0 (37.2-87.0) | 62.0 (36.8-86.2) | 80.0 (69.5-91.0) |
| Albumin level, g/L | 3.7 (3.3-4.1) | 4.1 (3.8-4.3) | 3.8 (3.2-3.9) | 3.7 (3.3-4.1) | 3.7 (3.3-4.2) | 3.6 (3.6-3.9) |
| ALT level, U/L | 23.0 (15.0-36.5) | 12.0 (7.0-23.0) | 25.0 (18.0-27.0) | 23.0 (16.0-36.0) | 23.0 (14.0-37.0) | 15.0 (15.0-22.0) |
| Total bilirubin level, mg/dL | 0.5 (0.3-0.7) | 0.3 (0.3-0.4) | 0.5 (0.3-1.2) | 0.5 (0.3-0.7) | 0.5 (0.3-0.8) | 0.6 (0.5-0.6) |
| CRP level, mg/L | 4.4 (1.4-11.6) | 1.8 (1.4-2.4) | 5.9 (3.1-11.7) | 5.8 (2.0-12.6) | 3.8 (1.1-11.0) | 4.2 (4.0-4.5) |
| IL-6 level, pg/mL | 40.0 (21.2-68.4) | - | 14.4 (14.4-14.4) | 42.4 (27.8-58.8) | 40.8 (19.9-94.3) | . (-.) |

SpO<sub>2</sub>/FiO<sub>2</sub> ratio, ratio of oxygen saturation by pulse oximetry to the fraction of inspired oxygen; GFR, glomerular filtration rate; ALT, alanine aminotransferase; CRP, C-reactive protein; IL-6, interleukin-6

**Supplementary Figure 3:**
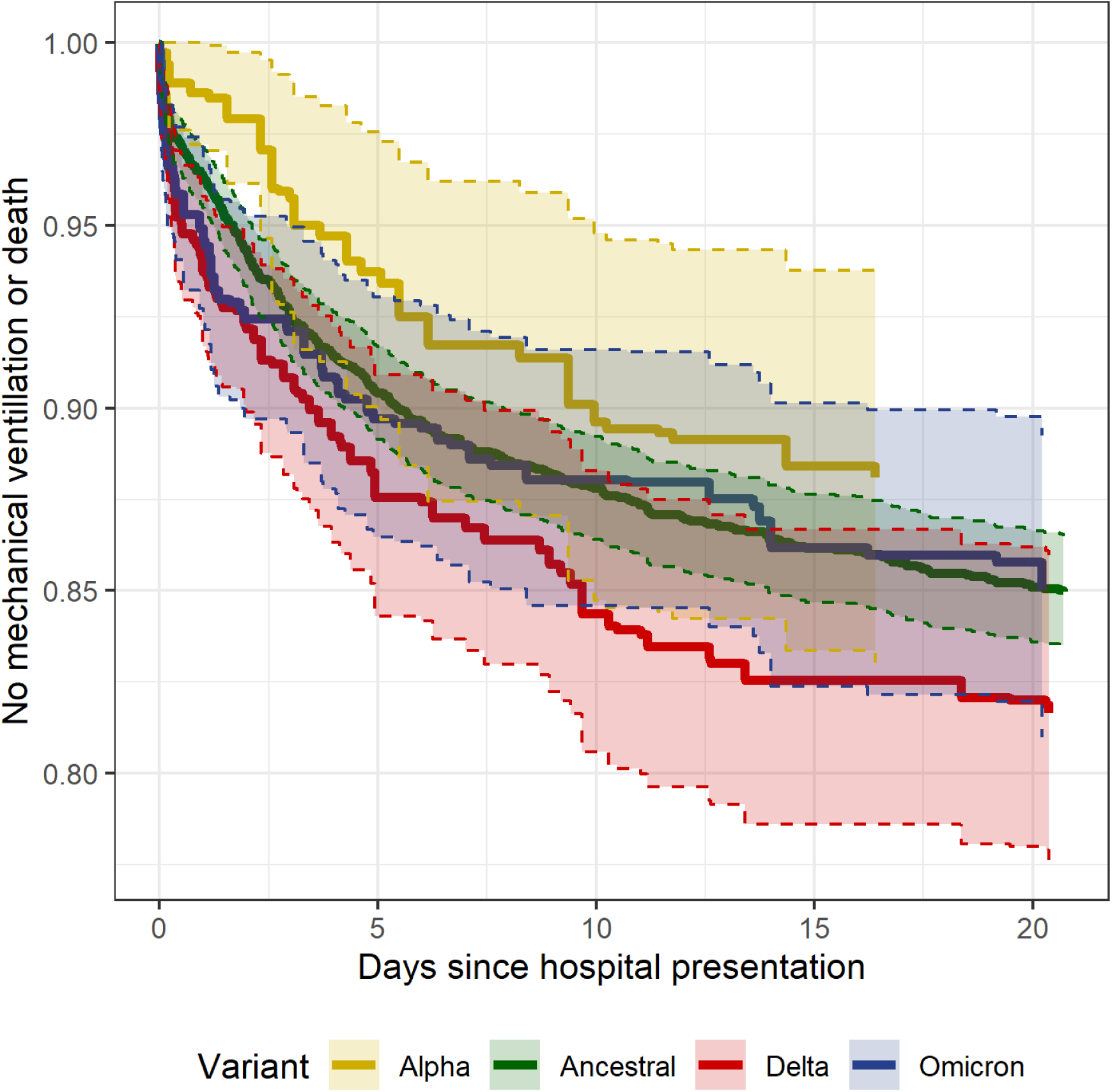
Inverse probability weighted risk of mechanical ventilation or death by days since hospital presentation and variant among unvaccinated patients with COVID-19.

**Supplementary Table 2.** Balance metrics for the average treatment effect model among unvaccinated patients

| Baseline characteristic used for propensity scoring | Ancestral | Alpha | Delta | Omicron |
| --- | --- | --- | --- | --- |
| Age, mean | 61.7475 | 60.8618 | 60.6444 | 61.4960 |
| Male, proportion | 0.5036 | 0.4821 | 0.5134 | 0.4963 |
| Elixhauser score, mean | 5.2050 | 5.2103 | 5.0573 | 5.1920 |
| Body mass index, kg/m<sup>2</sup> mean | 29.9679 | 29.8994 | 29.7269 | 29.7975 |
| Immunosuppressed or transplant patient, proportion | 0.0872 | 0.0997 | 0.0879 | 0.0792 |

**Supplementary Figure 4.**
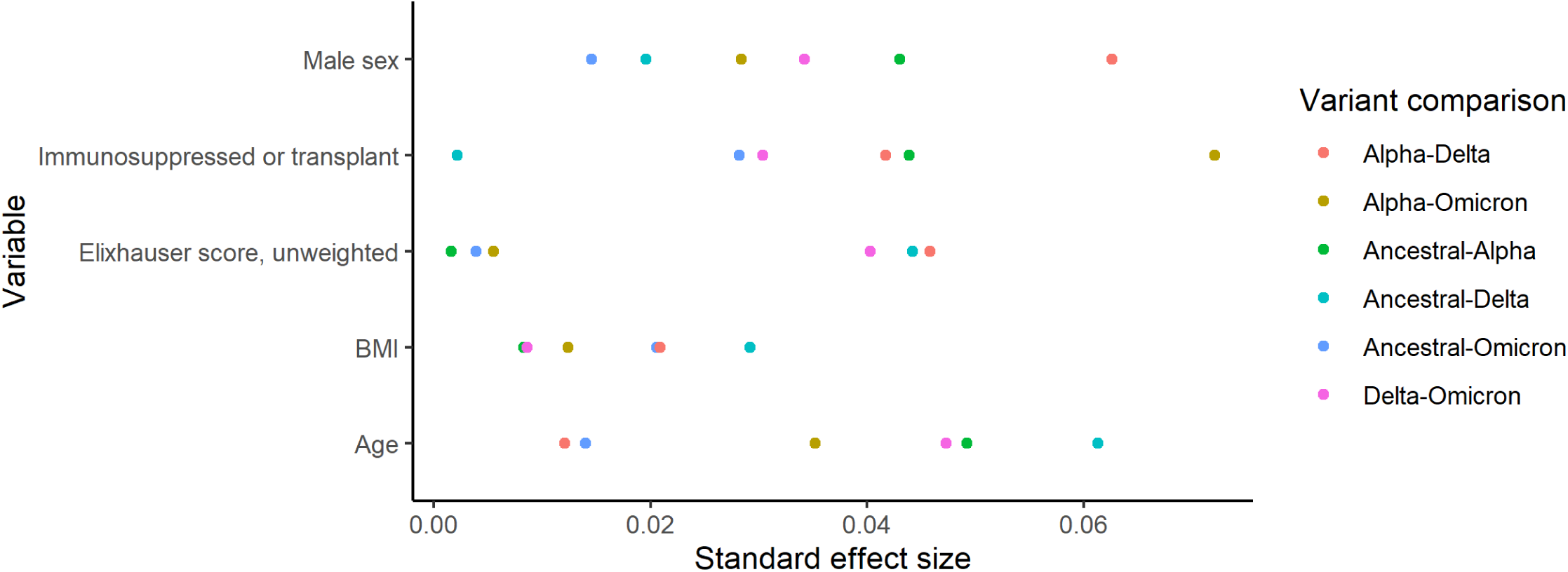
Standard effect size by variable used for propensity scoring and variant comparison for the average treatment effect model among unvaccinated patients

**Supplementary Table 3:** Clinical outcomes and features by variant in an average treatment effect model adjusted for risk factors for severe COVID-19, limited to 2,744 patients with inferred ancestral lineage or confirmed variant results

|  | Ancestral, n = 2744 | Alpha, n = 214 | Delta, n = 351 | Omicron, n = 63 |
| --- | --- | --- | --- | --- |
| Mean presenting SpO2/FiO2 | 362.48 (357.43, 367.53) | 368.57 (350.94, 386.21) | 341.92 (326.83, 357.02) | 375.14 (338.87, 411.42) |
| Mean Difference |  | 6.09 (-12.25, 24.44) | -20.56 (-36.47, -4.64) | 12.66 (-23.96, 49.29) |
| 14-day risk of severe disease or death |  |  |  |  |
| Estimate | 0.27 (0.25, 0.29) | 0.3 (0.23, 0.38) | 0.36 (0.3, 0.42) | 0.28 (0.15, 0.42) |
| Relative risk, Ancestral as reference | Ref | 1.13 (0.84, 1.42) | 1.33 (1.1, 1.56) | 1.06 (0.55, 1.56) |
| Relative risk, Delta as reference | 0.75 (0.63-0.89) | 0.85 (0.63-1.13) | Ref | 0.79 (0.48-1.3) |
| Risk difference, Ancestral as reference | Ref | 0.04 (-0.04, 0.11) | 0.09 (0.03, 0.15) | 0.02 (-0.12, 0.15) |
| Treatment with remdesivir |  |  |  |  |
| Estimate | 0.53 (0.5, 0.55) | 0.56 (0.47, 0.64) | 0.62 (0.56, 0.67) | 0.55 (0.4, 0.7) |
| Treatment with dexamethasone |  |  |  |  |
| Estimate | 0.72 (0.7, 0.74) | 0.77 (0.7, 0.83) | 0.75 (0.71, 0.8) | 0.57 (0.42, 0.72) |

**Supplementary Figure 5:**
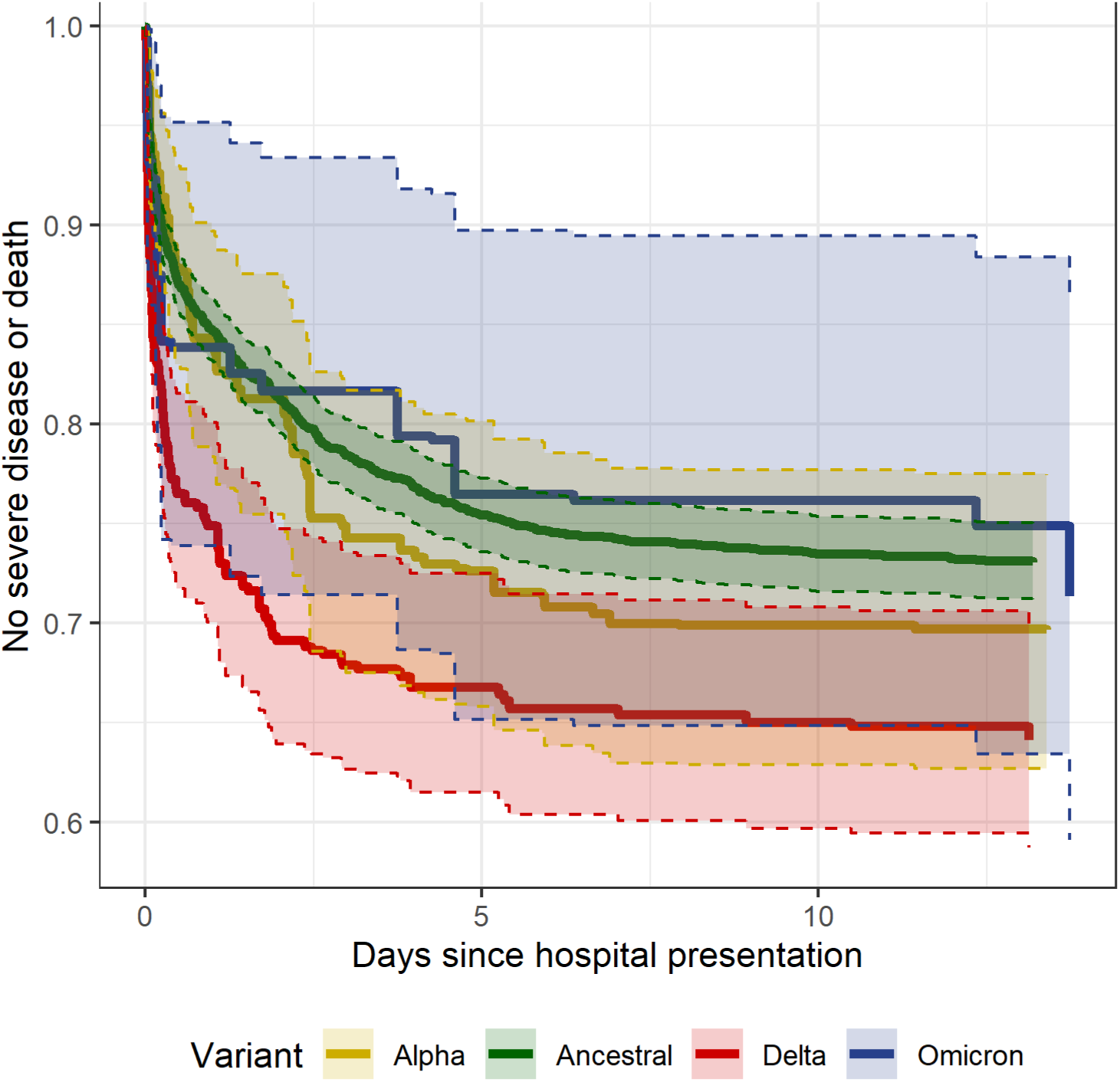
Sensitivity analysis of outcomes among unvaccinated persons limited to 2, 744 with inferred Ancestral lineage or confirmed variants, average treatment effect using inverse probability weighting

**Supplementary Table 4.**
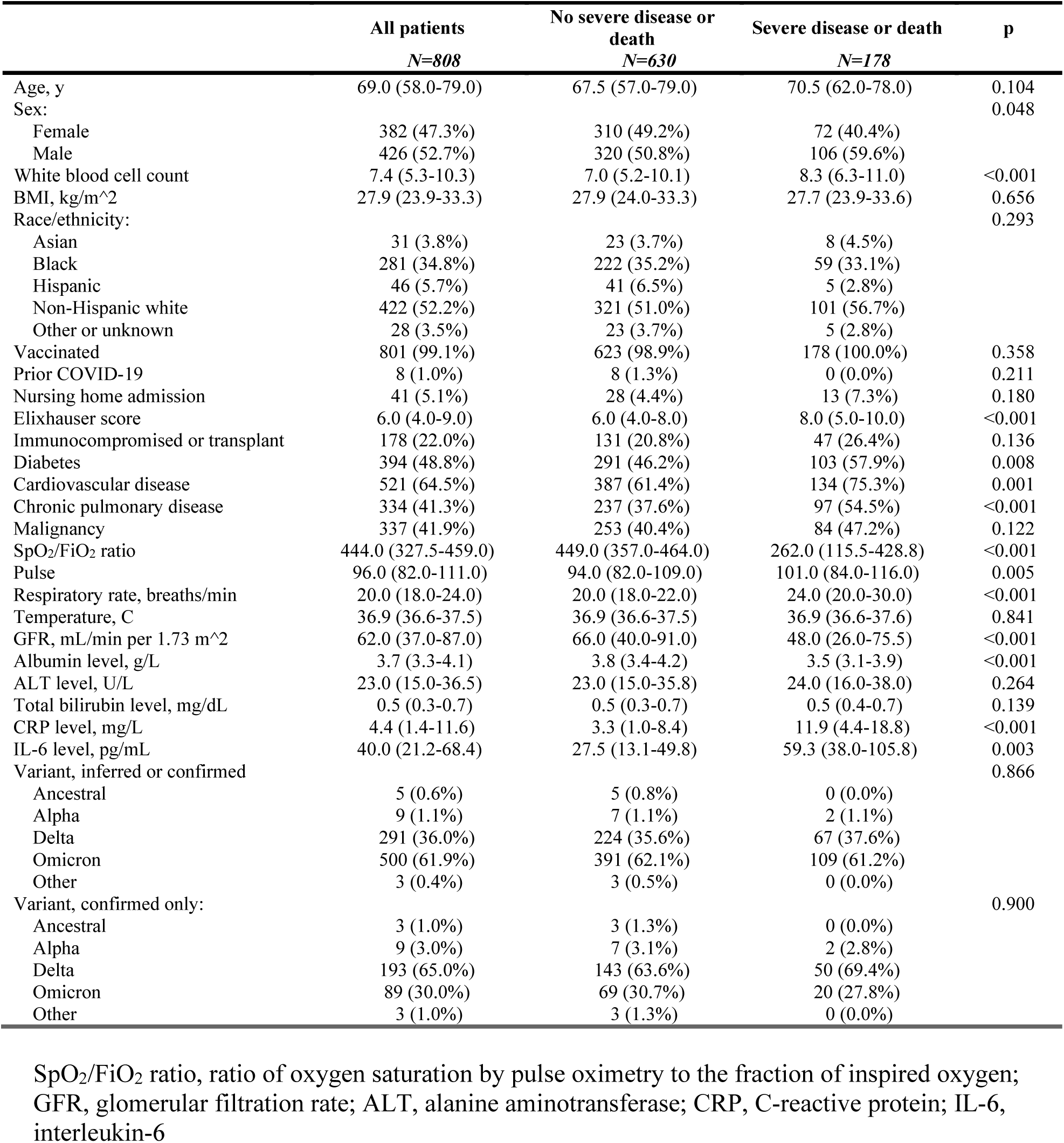
14-day outcome of severe disease or death among 773 hospitalized patients with history of vaccination or prior COVID-19

**Supplementary Table 5:** Severe disease or death within 14 days of hospitalization among 1,721 inpatients with confirmed or inferred Delta and Omicron variant infections

| **Clinical factor** | **Adjusted hazard ratio** | **p** |
| --- | --- | --- |
| Vaccinated or prior COVID-19 | 0.46 (0.34-0.62) | <0.001 |
| Age, by decade | 1.14 (1.07-1.21) | <0.001 |
| Elixhauser score | 1.08 (1.05-1.11) | <0.001 |
| Omicron variant | 0.71 (0.56-0.91) | 0.006 |
| Delta variant | Ref |  |
| Body mass index | 1.02 (1.01-1.03) | <0.001 |
| Immunosuppressed or transplant | 1.07 (0.82-1.4) | 0.632 |
| Interaction term: Omicron variant - vaccinated or prior COVID-19 | 1.35 (0.92-2) | 0.130 |

Adjusted hazard ratio reported for 457 events among 1,721 inpatients.

**Supplementary Figure 6:**
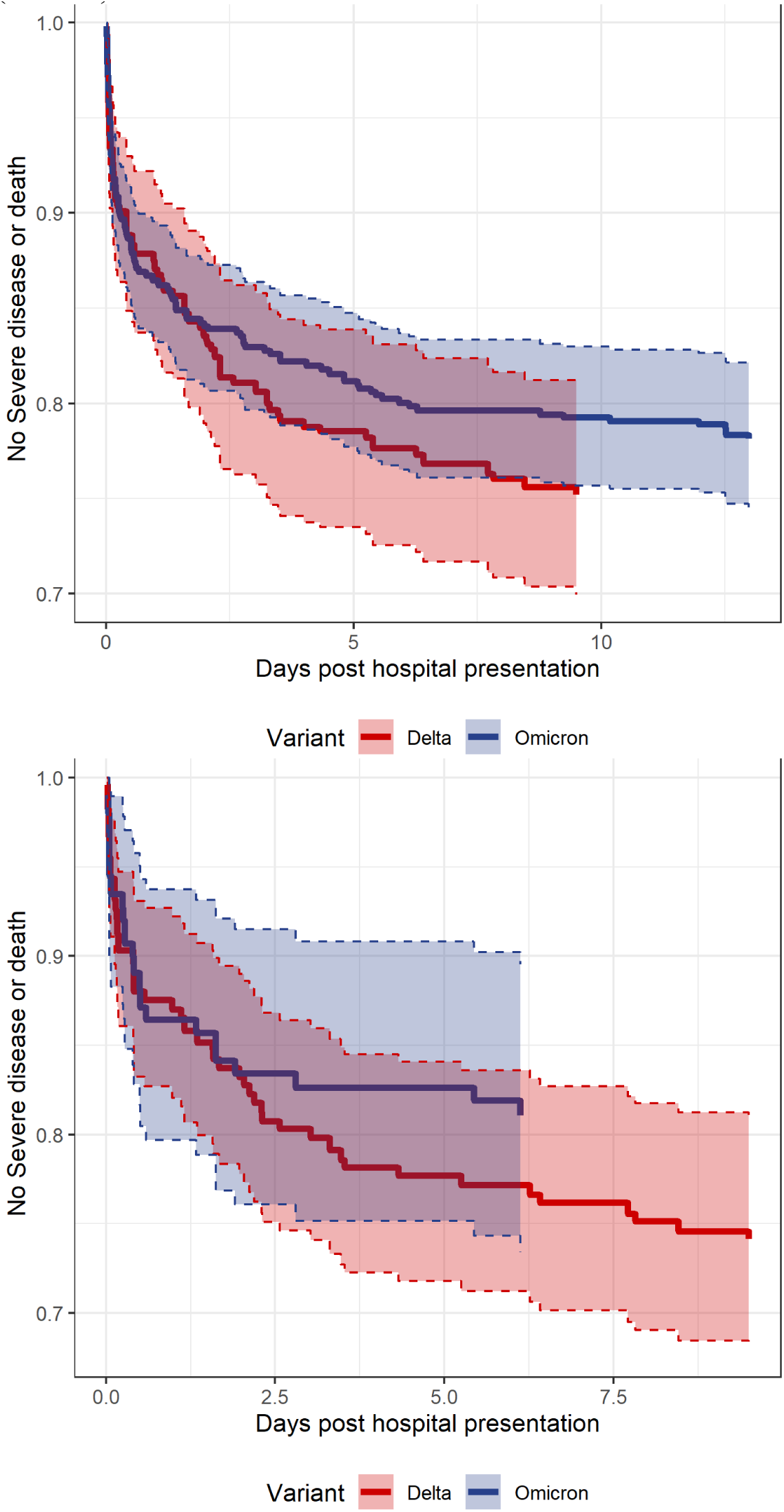
Inverse probability weighted risk of severe disease or death by days since hospital presentation and variant among vaccinated patients with inferred or confirmed Delta and Omicron infections (top) and limited to confirmed Delta and Omicron infections (bottom)

